# Multi-city modeling of epidemics using spatial networks: Application to 2019-nCov (COVID-19) coronavirus in India

**DOI:** 10.1101/2020.03.13.20035386

**Authors:** Bhalchandra S. Pujari, Snehal Shekatkar

## Abstract

The ongoing pandemic of 2019-nCov (COVID-19) coronavirus has made reliable epidemiological modeling an urgent necessity. Unfortunately, most of the existing models are either too fine-grained to be efficient or too coarse-grained to be reliable. Here we propose a computationally efficient hybrid approach that uses SIR model for individual cities which are in turn coupled via empirical transportation networks that facilitate migration among them. The treatment presented here differs from existing models in two crucial ways: first, self-consistent determination of coupling parameters so as to maintain the populations of individual cities, and second, the incorporation of distance dependent temporal delays in migration. We apply our model to Indian aviation as well as railway networks taking into account populations of more than 300 cities. Our results project that through the domestic transportation, the significant population is poised to be exposed within 90 days of the onset of epidemic. Thus, serious supervision of domestic transport networks is warranted even after restricting international migration.

## I. INTRODUCTION

Following its onset in Wuhan, China, the pandemic of 2019-nCov (COVID-19) coronavirus has wreaked havoc in various countries^1,2^. In less than three months the epidemic has engulfed over 100 nations with more than 100,000 confirmed infections and 4000 deaths^3,4^. Although the early cases indicate the illness is less severe than other coronaviruses like SARS-CoV and MERS-CoV, the evidences of rapid human-to-human transmission indicates that 2019-nCoV is more contagious than others^5^. Thus, it is utmost necessary to develop models that are computationally efficient yet realistic enough to assist medical personals, policy makers and general public.

One of the most celebrated models to study epidemics is the SIR model, and its subsequent variations^6–8^. This model distributes the total population into compartments for Susceptible, Infected and Recovered individuals, and a set of coupled differential equations describes the movement of population from one compartment to another. Although the model has been used extensively it fails to account for the demographic details and spatial heterogeneity. Several remedies have been proposed to overcome this limitation. For example, ‘fully-mixed’ assumption can be replaced with network of contacts between individuals^9,10^. In the last few days network based SIR-type models were also used to predict the spread of ongoing epidemic of COVID-19 coronavirus^11,12^. Many studies have used the transmission dynamics of virus in metapopulation patches where the total population is subdivided into a number of discrete patches, each of which is treated as being well-mixed^13^. Bolker et al.^14^ used such a model to demonstrate the effectiveness of vaccination against measles outbreak. Arino and van den Driessche^15,16^ have proposed a model in which the population of a city is coupled with other cities via a mobility parameter that describes the net rate of intercity migration^16^. Lloyd and Jansen^17^ also used meta-population of *n*-*patches* to epidemics’ dynamics to see the patterns of synchrony in outbreaks. A more involved model with cross-patch infection was investigated for various cases by Muroya et al.^18^, who looked at the global stability of an endemic.

We note that the network based models are computationally inefficient while those using metapopulations, although efficient, usually fail to maintain initial populations of patches. In fact Arino and van den Driessche^15^ have pointed out that a multi-city model achieves an equilibrium population that may not be the one started with.

Here we introduce a new technique to main-tain the desired population of a city via self-consistent parametrisation. In this approach, disease dynamics inside individual cities are treated with SIR model that accounts for the intra-city mixing. On the other hand, intercity migration is modeled by spatially realistic networks of airports and train stations. Put differently, the “fully-mixed” approximation is assumed to be valid inside cities but not between the cities. Another notable feature of the scheme proposed here is the introduction of distance-dependent delay inherent in the transportation. Additionally, the model is also capable of simultaneously handling cities with different populations. This is achieved by scaling the product terms inside the SIR equations by respective populations as discussed in the next section. As an application to ongoing COVID-19 outbreak, we run our model on aviation and rail networks of India where links connect multiple cities of varying population sizes.

The paper is organised as follows: In Sec. II, we outline the details of our model. Then in Sec. III we discuss our main findings in depth. In the last section, Sec IV we summarise the results and their implications.

### II. METHODOLOGY

We assume the existence of several metapopulation units inside a given geographical region such that for each of these the dynamics of disease is well approximated by the standard SIR model. The disease transmission among the units is due to migrating population described using a coupling term.

In classic SIR model the *s, x*, and *r* represent the susceptible, infected and recovered fractions respectively. However while dealing with multiple meta-populations of heterogeneous sizes we are required to represent them in arbitrary units, typically taking smallest meta-population as the unit. If *ϕ*_*i*_ = *s*_*i*_ + *x*_*i*_ + *r*_*i*_ is the size of metapopulation *i* in these units, then product terms in SIR model must be divided by *ϕ*_*i*_ for sake of consistency.

We can view this system as a network in which metapopulations are the nodes that are connected by links representing migration. We assume that the outward migration rate from node *i* is proportional to its size *ϕ*_*i*_. Assuming that a migrating individual at *i* is equally likely to migrate to any of the *k*_*i*_ neighbours, the fraction of population reaching a neighbour is proportional to 1*/k*_*i*_.

Finally, we note that, in reality when dealing with spatially extended network, migration between *i* and *j* takes place with finite speed and hence there is an associated delay *δ*_*ij*_, which we consider to be proportional to the geographic distance between *i* and *j*. Combining all these, our model of interacting meta-populations is the following

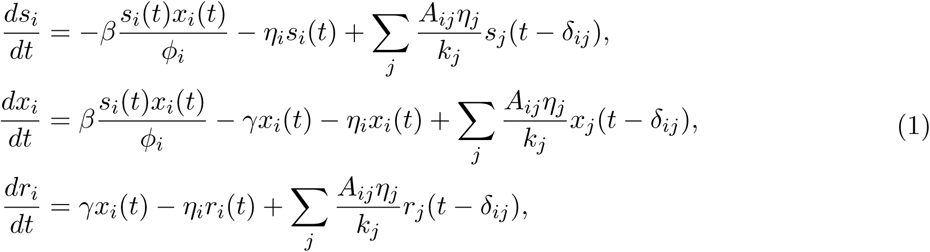

where *A* is the adjacency matrix of the network with *A*_*ij*_ = 1 if *i* and *j* are connected, and 0 otherwise. The *β*, and *γ* are respectively the infection and transmission rates as used in classic SIR model, with *basic reproduction number R*_0_ = *β/γ*. Here we choose *β* = 0.2 and *γ* = 0.07 (per day) for COVID-19 following the recent estimate by Zhu et al^11^.

Now we make a crucial observation that in the real world, in spite of migrations the total population of a city remains more or less constant during the span of a typical epidemic. To enforce this condition the net migration of a node should be zero, although the same cannot be said for *s, x*, and *r* sub-populations. Thus

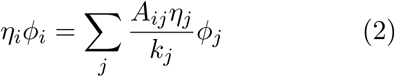

This is a self-consistent equation for *η*_*i*_’s, and can be solved iteratively, starting from arbitrary values of *η*’s. The *η*’s thus obtained are determined up to a proportionality constant, say *D*.

In the subsequent sections we consider India to be our geographical region and cities as meta-population units. The required data is obtained from various online resources^19–21^.

## III. RESULTS AND DISCUSSION

We apply our model to study the spread of ongoing epidemic of 2019-nCov (COVID-19) in India using aviation and railway networks as substrates. In these networks a link represents a direct flight/train between the given nodes. The sizes of our aviation and train networks are 69 and 320 respectively. Given the time scale of epidemic, it is reasonable to assume that the transport from city *i* to *j* via air is instantaneous, making the delay *δ*_*ij*_ = 0. However the same assumption cannot be made for train transport, and we make the corresponding delay *δ*_*ij*_ proportional to the geographic distance between the stations by assuming the average speed to be 50 km/h.

As mentioned above, the *η*’s are determined only up to the scale *D* by Eq(2), and hence their converged values depend on the initial guess. We start by calculating *η*_*i*_’s with starting guess of *η* = 1 for all the nodes. In practice, *D* depends on the actual migration data and its accurate estimation is beyond the scope of this work. Instead, we vary *D* and examine the behaviour of the system.

We investigate the infected population of all the cities in time for both the networks separately, by first infecting Delhi with *x*^Delhi^(0) = 0.0001*ϕ*^Delhi^ as it is well connected internationally. Fig. 1 shows the time series of infected population of several cities for *D* = 0.01. We note several key features of these time series. First, as a result of our modification of the SIR model that allows us to handle heterogeneous population sizes, the values of the maxima are proportional to the populations of the corresponding cities. Second, because *β* and *γ* are same for all the cities, bigger cities take longer to achieve their maxima. Also, for both the networks, most of the maxima occur in close proximity with each other implying the necessity of the preparedness against simultaneous large-scale outbreaks. Since the train network includes delay, the maxima tend to occur much later relative to that of Delhi. Because the train transport is the dominant means of transport in India, this shift of the peaks is more relevant for the response against coronavirus.

**FIG. 1:**
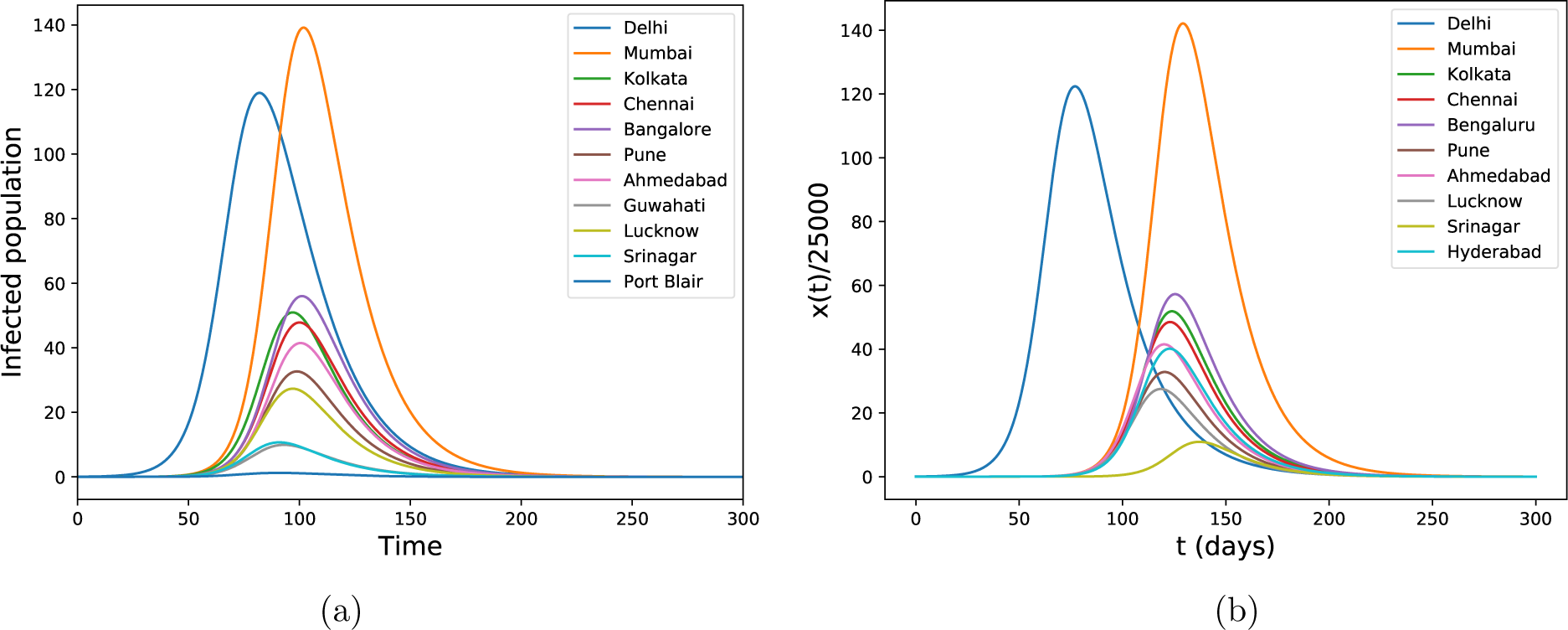
The time series of the infected populations for select cities of (a) the aviation network, and (b) the train network of India when Delhi is infected initially. Both the values of the maximum and its location are governed by the population of the corresponding city

After this, We take a more realistic situation in which more than one city is initially infected. For this, we consider six different cities and corresponding initial infected fractions: Delhi (10^*−*4^), Mumbai (10^*−*5^), Chennai (10^*−*5^), Kolkata (2*×* 10^*−*5^), Bengaluru (10^*−*5^), and Kochi (10^*−*6^) with *D* = 0.01. The results are shown in Fig. 2 for the train network. The cities which are initially infected tend to peak earlier hence the peaks are no longer as synchronous as before. Because the delay in the air network is zero, this loss of synchrony is not as prominent, and we show only the network snapshots in Fig. 3. From the figure, we see that almost all the nodes become maximally infected after about 90 days.

**FIG. 2:**
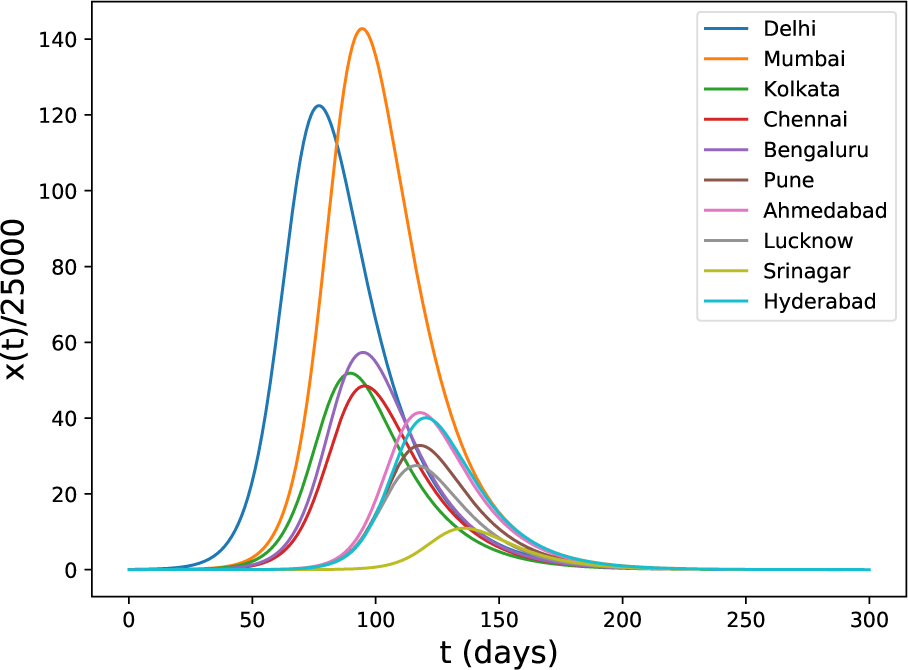
The time series of infected population when multiple cities are initially infected. Delhi, Mumbai, Kolkata, Chennai are Bengaluru are initially infected.

**FIG. 3:**
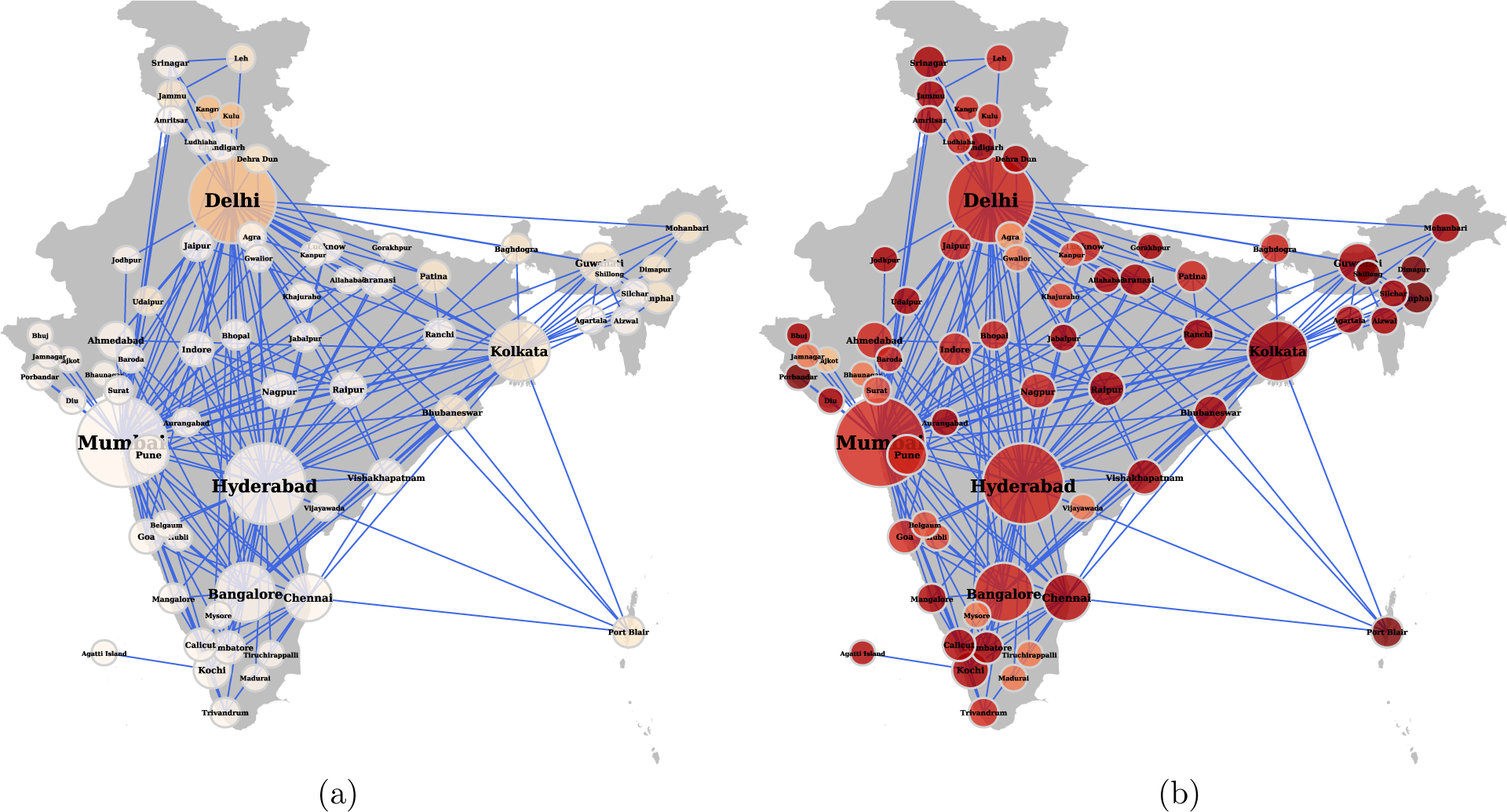
Snapshots of the Indian aviation networks after 60 days (left) and after 90 days (right) when several internationally well connected cities are infected. The dark red color indicates higher levels of infected fractions of populations. After 60 days, a sudeen growth in the infected population is clearly seen.

The accurate estimation of the parameter *D* which is related to the migration is unavailable. Thus, to check its effect on the epidemic, we vary it and observe the behaviour of the time *t*_max_ at which the infection maximum occurs for each city. The results are shown in Fig 4 with initial infection only at Delhi. *D* = 0 corresponds to quarantining Delhi from the rest of the nodes, and hence only Delhi has finite *t*_max_ while for others *t*_max_ is undefined. For *D >* 0, infection spreads to other nodes, and understandably very small values of *D* result in very large values of *t*_max_. Upon increasing *D*, the value of *t*_max_ decreases rapidly as more fraction of individuals migrate among the cities. Interestingly, for each node except the initially infected one, there exists a critical value of *D* at which this trend reverses and *t*_max_ starts increasing again. We can understand this unexpected behaviour with the following argument. When *D* is very high, it is analogous to having all the population well-mixed. As a result, too many infected individuals migrate from node to node to infect any one node effectively. In other words, too few infected individuals remain in any given city compared to low *D*. As a result, even the initially infected node takes long time to get maximally infected, and peaks for all the cities appear simultaneously.

**FIG. 4:**
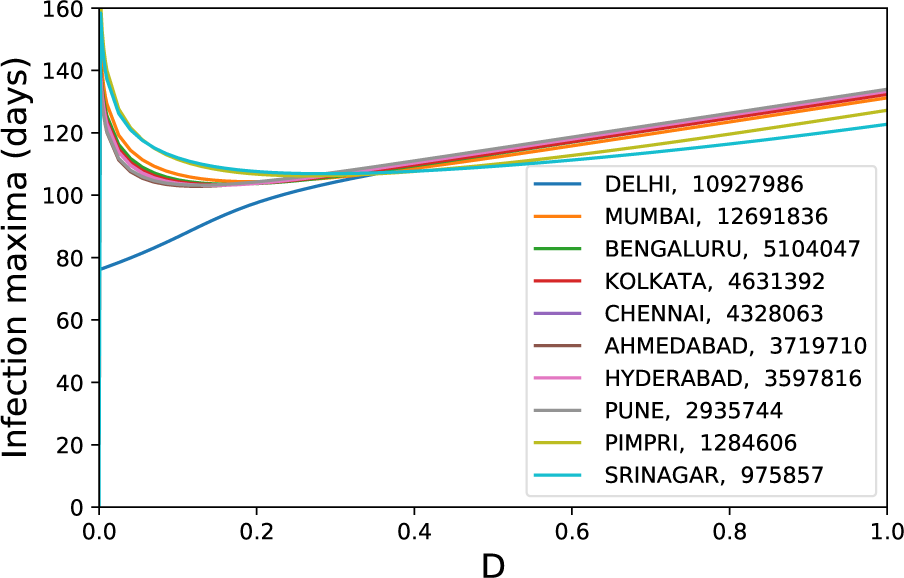
The variation of *t*_max_ with *D* for several cities, when only Delhi is infected initially. Legend also shows the populations of the corresponding cities.

## IV. CONCLUSION

Although the spread of epidemic on spatially extended systems is a well studied area, the majority of models are either computationally expensive since they consider individuals and interactions between them, or too coarse-grained to account for spatial migrations. Here we have proposed a novel approach that addresses both these issues, and offers fast and realistic predictions that are useful for policy-makers and health personnels. Our hybrid approach to SIR model with well-mixed intra-city populations and intercity coupling based on transportation network lets us predict the course of ongoing pandemic of COVID19 coronavirus in India. The forecasts based on our model indicate that within 90 days of the start of the epidemic, most of the urban population will be exposed to the virus unless preventive measures are taken. Our work also shows that because of the contagious nature of the COVID19 and the crucial role of the domestic transportation networks, even a small infected population is sufficient to sustain and spread the pandemic. Thus, along with restricting international links, it is equally important to monitor the domestic transportation.

## Data Availability

The data used in the preprint is taken from the following three places, and is public. LInks to the data are given in the manuscript. https://openflights.org/

https://openflights.org/

https://github.com/datameet/railways

https://worldpopulationreview.com/countries/india-population/cities/

## Acknowledgements

SMS. acknowledges the funding from the DST-INSPIRE Faculty Fellowship (DST/INSPIRE/04/2018/002664). BSP acknowledges support from UGC Faculty Recharge Programme. Some analysis was carried out using *graph-tool*^22^.

